# High-resolution disconnectome predicts outcome and response to thrombectomy in basilar artery occlusion

**DOI:** 10.64898/2026.04.20.26350998

**Authors:** Benjamin Authamayou, Gaultier Marnat, Anna Matsulevits, Fanny Munsch, Audrey Lavielle, Nadège Corbin, Chris Foulon, Bailiang Chen, Emilien Micard, Benjamin Gory, Vincent L’Allinec, Romain Bourcier, Olivier Naggara, Eliot Lauze, Grégoire Boulouis, Bertrand Lapergue, Omer Eker, Igor Sibon, Michel Thiebaut de Schotten, Thomas Tourdias, ETIS investigators

## Abstract

**Background:** Acute basilar artery occlusion (BAO) causes devastating strokes. Despite the benefit of endovascular treatment, the optimal management remains sometimes controversial, such as for patients with mild deficits, and would benefit from robust prognostic tools. Given the dense white matter networks within the posterior fossa, we tested whether quantifying disconnections from acute diffusion-weighted imaging (DWI) could improve outcome prediction and responders to recanalization compared with conventional metrics.

**Methods:** We conducted a secondary analysis from a prospective multicenter stroke registry, including consecutive patients (2017–2024) with BAO and admission MRI. Ultra-high-resolution diffusion MRI was acquired in healthy participants to build normative tractograms with optimized posterior fossa quality. Patient infarcts delineated on DWI were projected onto these tractograms to estimate disconnected fiber volume. The primary outcome was 90-day modified Rankin Scale (mRS) 0–3 vs 4–6. Predictive performance of disconnected fiber volume was compared with baseline NIHSS, infarct volume, and posterior circulation ASPECTS (pc-ASPECTS) using logistic regressions and areas under receiver operating characteristic curves (AUC). Ordinal regressions tested associations across the full mRS spectrum, stratified by recanalization status. Analyses were repeated in patients with NIHSS ≤10.

**Results:** Among 201 patients (median age 70; NIHSS 10), 97 (48.3%) had poor outcome. Despite small median infarct volume (4.75 mL), disconnected fiber volume was substantial (median 25.15 mL). Disconnected fiber volume achieved an AUC of 0.84, outperforming NIHSS (0.67; p<0.0001), infarct volume (0.75; p=0.00059), and pc-ASPECTS (0.76; p=0.0127). Low disconnected fiber volume predicted better outcomes across the full mRS (OR=0.12 [95% CI, 0.065-0.204]) and greater benefit from successful recanalization (OR=0.33 [95% CI, 0.15-0.70]). In patients with NIHSS ≤10 (n=102), disconnected fiber volume remained the strongest predictor (AUC=0.83).

**Conclusions:** Disconnected fiber volume derived indirectly is a robust prognostic marker of BAO outcomes that outperforms conventional predictors and may support future treatment decisions.

**Registration:** https://clinicaltrials.gov - NCT03776877.

## Introduction

Acute basilar artery occlusion (BAO) accounts for only ∼1% of ischemic strokes yet is among the most devastating, with high risks of severe disability and mortality ^1^. Endovascular treatment (EVT) is now an established therapeutic option for BAO within 24 hours of symptom onset ^2, 3^. Despite this, outcomes remain poorer than for anterior circulation strokes, and treatment decisions are often complex ^4^. Futile recanalization raises major ethical challenges as it could prevent death while leaving patients with severe disability. It also carries significant iatrogenic risks and imposes substantial healthcare costs. In particular, the optimal management of patients presenting with milder symptoms (National Institutes of Health Stroke Scale, NIHSS <10), whether to perform EVT, administer thrombolysis, or opt for close monitoring without immediate reperfusion, remains controversial ^4, 5^. These challenges highlight the pressing need for robust prognostic stratification tools to guide individualized therapeutic decisions.

While some innovative deep learning approaches are starting to emerge for the prediction of thrombectomy outcomes ^6^, these models are limited to anterior circulation strokes, a totally distinct clinical scenario. The current predictors of long-term functional outcome in BAO remain suboptimal. The NIHSS, although widely used in clinical practice, is poorly suited to posterior circulation strokes ^7^, leading to the development of specific posterior circulation stroke scales ^8^. Imaging-based tools such as the posterior circulation Alberta Stroke Program Early CT Score (pc-ASPECTS) adapt the original ASPECTS for this territory ^9^, providing a semi-quantitative estimate of infarct extent and location. Nevertheless, pc-ASPECTS has demonstrated at best moderate prognostic accuracy for long-term disability ^10, 11^. A key limitation of these tools is their reliance on traditional topographical approaches that treat brain regions as isolated functional units.

Notwithstanding, neurological deficits and recovery potential depend largely on the integrity of distributed brain networks ^12^. Disruption of these connections, so-called “disconnection”, is often a stronger determinant of symptoms than lesion location alone ^12–14^. The posterior fossa contains dense white matter networks essential for motor, sensory, arousal, and swallowing functions. Even small strategically located lesions can cause disproportionately severe deficits by disconnecting these critical pathways ^15^.

Rapid assessment of brain connectivity and disconnectivity could therefore provide valuable prognostic information in BAO. Still, capturing such information in routine clinical practice remains a cutting-edge challenge. In recent years, a pragmatic indirect method has emerged ^16^: projecting individual stroke lesions onto normative tractograms from healthy participants to infer probable white matter disconnections, even when patient-specific high-angular, multi-shell diffusion weighted imaging is unavailable, as it is the case in acute stroke. In anterior circulation stroke, this “disconnectome” approach has shown promise in predicting both cognitive and functional outcomes ^14, 17, 18^. Yet, its application to BAO has not been specifically investigated, partly due to additional methodological challenges related to spatial resolution ^19^ and cardiac artifacts ^20, 21^, among others.

In this study, we optimized a high-resolution disconnectome-based framework using acute MRI to accurately estimate white matter disconnections caused by BAO. We hypothesized that the extent of fiber disconnection would improve prediction of functional outcomes and help identify patients most likely to benefit from EVT versus those for whom the intervention would likely be futile, compared with conventional topographical and volumetric imaging metrics.

## Materials and Methods

### Study Population

We conducted a secondary analysis of the Endovascular Treatment in Ischemic Stroke (ETIS; ClinicalTrials Identifier: NCT03776877, approved by the ethical committee n° 2018-09 under ID-RCB n°: 2017-A03457-46) registry, an ongoing, prospective, multicenter, real-world observational registry evaluating large-vessel occlusion in 21 comprehensive stroke centers in France. Clinical data and brain imaging are provided by participating centers and stored centrally. Written informed consent was obtained from all patients or their legal representatives. This analysis, named BAO-DISCO (Basilar Artery Occlusion Disconnectivity), is reported according to the Strengthening the Reporting of Observational Studies in Epidemiology (STROBE) guidelines for observational studies ^22^.

From the ETIS database, we included all consecutive patients from 2017 to 2024 who met the following criteria:

1. Age ≥18 years,
2. Acute BAO, either isolated or associated with intracranial vertebral artery or posterior cerebral artery occlusion, confirmed by baseline vascular imaging,
3. Pre-treatment brain MRI including a diffusion-weighted imaging (DWI) sequence.

### Data collection

From the ETIS electronic case report form, we extracted the following variables: age, sex, baseline NIHSS score, stroke etiology, vascular risk factors, time from symptom onset to acute MRI, treatment type (intravenous thrombolysis and/or EVT), time from onset to recanalization, and recanalization success defined as Thrombolysis in Cerebral Infarction (TICI) ≥2b.

The primary outcome was the modified Rankin Scale (mRS) score at 3 months collected by certified investigators during routinely scheduled visits or by trained research nurses during a standardized telephone interview. A good outcome was defined as mRS 0–3 for ambulatory patients, and a poor outcome otherwise, consistent with primary outcome definitions used in clinical trials of EVT for BAO ^2, 3, 5, 23^.

### Imaging Analysis

We analyzed acute DWI and the corresponding apparent diffusion coefficient (ADC) maps centrally, blinded to clinical data. Images were acquired at either 1.5T or 3T, with acquisition parameters that could vary across centers: repetition time ranged from 2800 to 8400 ms, echo time from 54 to 102 ms, acquisition matrix from 256×256 to 384×384, and b-values of 0 and 1000 s/mm² (1500 or 2000 s/mm² in a few cases).

Infarcts were manually delineated to determine stroke volumes prior to any treatment and to infer disconnected white matter fibers (see below). Given the imperfect accuracy of automated lesion segmentation in the posterior fossa, delineation was performed manually by a first reader (BA) identifying areas with hyperintense DWI and low ADC, blinded to outcomes, and reviewed by a second senior reader (TT) for corrections.

Each individual DWI and the corresponding infarct mask were co-registered to the Montreal Neurological Institute (MNI152) standard space. We first tested the combination of rigid, affine, and non-rigid transformations with ANTs - advanced normalization tools ^24^. However, the brainstem on DWI is prone to geometric distortions in the phase-encoding direction due to adjacent air-filled structures, frequently leading to imperfect MNI alignment even after non-rigid registration (**Supplementary-Figure-1-A,B,C**). Therefore, in a second step, we created an intermediate study-specific DWI template by co-registering all DWI scans together. Subsequently, ANTs was employed to register each individual DWI scan to our optimized template. Then this template was adjusted to MNI152 using linear (FLIRT as part of the FSL package ^25^) and manual corrections (MIPAV v11.3.3; https://mipav.cit.nih.gov) to finely optimize anatomical alignment (**Supplementary-Figure-1-D**). Normalization quality was visually inspected by senior neuroradiologist (TT). A prevalence map was generated from co-registered infarct masks to explore the spatial distribution of infarcts.

### Disconnectome Analysis

To estimate fiber disconnection, we used an indirect approach in which normalized infarct masks were projected onto whole-brain tractography available in MNI152 space, as previously described for supratentorial stroke ^14, 16, 17^. However, resolving the complex fiber architecture of the brainstem and posterior fossa requires exceptionally high-quality data. We therefore specifically acquired new ultra–high-resolution diffusion tensor imaging datasets from healthy participants. In this context, we prioritized the quality of the connectome rather than age matching with the stroke cohort, since the shape and spatial extent of average tracts ^26^, as well as disconnection estimates ^16^, have been shown to remain invariant across decades. Moreover, in a previous indirect disconnectome study of supratentorial stroke, we have already demonstrated that age of the healthy participants used to estimate disconnections was not a confounding factor ^17^.

Accordingly, we collected very long scans (approximately 10 hours each) for one male participant (age range 40-50 years) and one female participant (age range 30-40 years) to reconstruct the highest-quality tractograms. For each patient, infarct masks were then applied to filter these tractograms, retaining only fibers intersecting the lesion. The intersection of the two filtered tractograms was taken as the set of streamlines consistently disconnected across both high-resolution datasets, from which we quantified the total volume of disconnected fibers for each patient (**Figure-1**).

**Figure 1:**
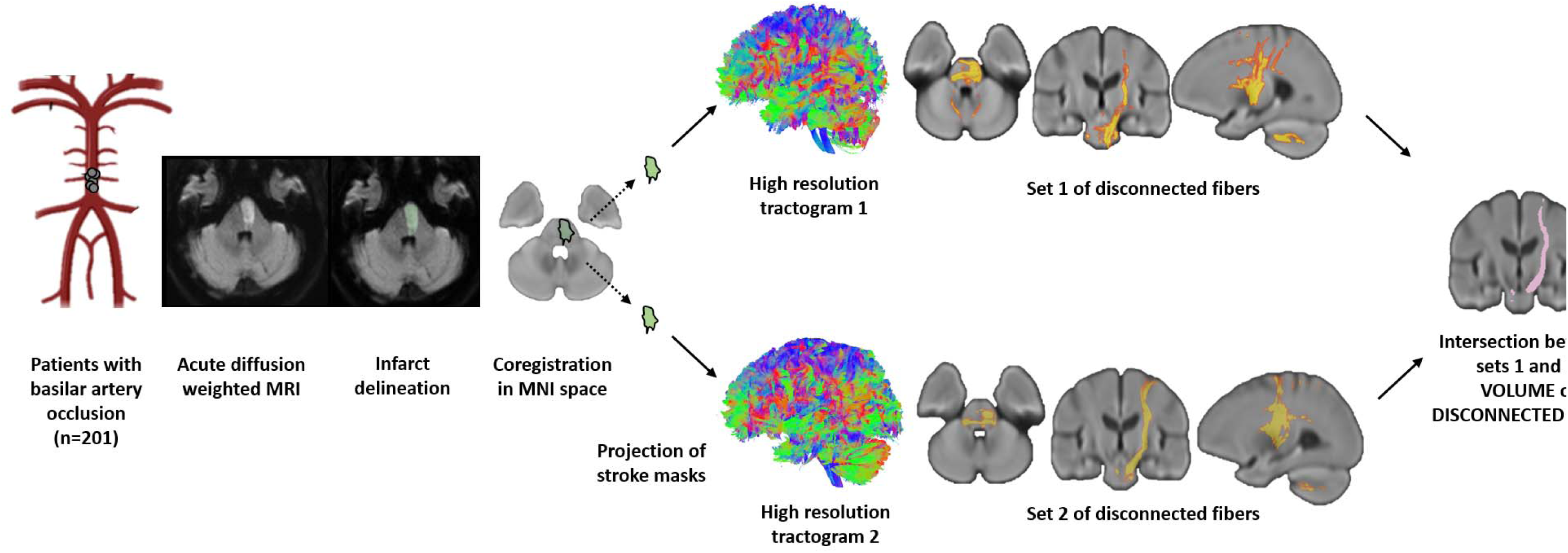
Disconnected fiber volume pipeline. Illustrative pipeline used to compute indirectly the volume of disconnected fibers associated with each basilar artery occlusion stroke.

The details of image acquisition and reconstruction for the high quality tractograms from the healthy participants were the following. The images have been collected on a Siemens 3T Prisma scanner (Siemens Healthineers, Erlangen, Germany). We chose the readout-segmented echo-planar imaging sequence (RESOLVE) ^27^ with cardiac triggering and 2D navigator controlling in real-time the reacquisition of unusable data to mitigate both image distortions and cardiac induced artefacts. Participants underwent five repetitions of the RESOLVE acquisition including 32 diffusion encoding directions with b-value of 2000 s/mm² for the four first sessions and 1000 s/mm² for the last session. Additionally, two acquisitions with null b-values were obtained with opposite phase encoding direction to subsequently correct for residual distortion. Acquisition parameters were: 2D spin-echo single-shot echo planar imaging with 1 mm isotropic resolution, repetition time 690 ms, echo times 81 ms and 132 ms, field-of view 220×220×128 mm^3^, 5 readout segments, GRAPPA acceleration factor 2.

The acquisition time of a single session was approximately 2 hours depending on the heart rate of the participant, leading to about 10 hours of acquisition in total per participant for the 5 sessions that were merged. We also collected a 3D T1 weighted acquisition with inversion time 880 ms, isotropic resolution of 1mm, TR/TE 2000 ms / 2.05 ms, flip angle 8°, field of view 256×256×192 mm^3^, and GRAPPA acceleration factor of 2.

Subsequently, susceptibility-induced off-resonance field was estimated from pairs of b0 with opposite phase of encoding direction ^28^ and corrected for the whole diffusion-weighted dataset using TOPUP ^29^. Motion and geometrical distortion were corrected using the EDDY tool as implemented in FSL. Next, StarTrack software (https://www.mr-startrack.com) calculated a damped Richardson-Lucy algorithm for spherical deconvolution and deterministic tractography in the native DWI space. A fixed fiber response corresponding to a shape factor of α = 1.5 × 10^−3^ mm^2^ s^−1^ was adopted, coupled with the geometric damping parameter of 8. Four hundred algorithm iterations were run. The absolute threshold was defined as three times the spherical fiber orientation distribution (FOD) of a grey matter isotropic voxel and the relative threshold as 8% of the maximum amplitude of the FOD ^30^. A modified Euler algorithm ^31^ was used to perform the whole brain streamline tractography, with an angle threshold of 35°, a step size of 0.5 mm and a minimum streamline length of 15 mm. We co-registered these structural connectome data to the standard MNI 2 mm space using the following steps: first, whole brain streamline tractography was converted into streamline density volumes where the intensities corresponded to the number of streamlines crossing each voxel. Second, a study-specific template of streamline density volumes was generated using the Greedy symmetric diffeomorphic normalisation (GreedySyN) pipeline distributed with ANTs ^24^. This provided an average template of the streamline density volumes for all subjects. The template was then co-registered with a standard 2 mm MNI152 template using flirt tool implemented in FSL ^25^. This step produced a streamline density template in the MNI152 space. Third, individual streamline density volumes were registered to the streamline density template in the MNI152 space template, and the same transformation was applied to the individual whole-brain streamline tractography using the trackmath tool distributed with the software package Tract Querier using ANTs GreedySyn ^32^. This step produced a whole brain streamline tractography in the standard MNI152 space.

### Statistical Analyses

Continuous variables were reported as medians with interquartile ranges; categorical variables as counts and percentages. Between-group comparisons (good vs. poor outcome) were performed using Student’s *t*-test, Mann–Whitney *U* test, or χ² test, as appropriate.

Variables differing significantly between outcome groups were entered into logistic regression models. Prognostic performance was evaluated using the area under the receiver operating characteristic curve (AUC) with 95% confidence intervals (CIs) obtained from 1,000 bootstrap replications. AUCs were compared using DeLong’s test. Predictive performance was further assessed with classification tables comparing predicted vs. observed outcomes.

The relationship between the volume disconnected fibers and other imaging metrics (infarct volume, pc-ASPECTS) was explored using linear regression.

To assess the effect of disconnected fiber volume across the full mRS range, we also performed a shift analysis: disconnected fiber volume was dichotomized as low vs. high using the Youden index ^33^ to determine the optimal threshold, then a proportional odds ordinal regression was run with 3-month mRS (0–6) as the outcome and disconnected fiber volume group as the predictor. Ordinal regression was repeated within each fiber volume subgroup (low and high) to assess whether the effect of successful recanalization on mRS shift varied by baseline disconnection severity.

Given the uncertainty surrounding treatment strategies for patients with mild deficits despite BAO ^4^, we repeated the previous analyses in the subgroup of patients with baseline NIHSS ≤10.

Analyses were conducted in R (version 4.0.3) and GraphPad Prism (version 10.4.1). Two-tailed *p* values <0.05 were considered statistically significant.

## Results

### Baseline Characteristics

From 2017 to 2024, 219 patients in the ETIS registry presented with BAO and underwent MRI before treatment. Eighteen were excluded (14 with missing MRI data, 1 lost to follow-up without mRS assessment, 1 withdrew consent, 1 reclassified as no BAO after secondary review, and 1 with concomitant middle cerebral artery stroke), leaving 201 patients with complete clinical and MRI data for analysis.

Baseline characteristics are shown in **Table 1**. The median age was 70 years (IQR, 58-77), 59.2% were male, and the median baseline NIHSS was 10 (IQR, 6-18). Intravenous thrombolysis was administered to 41.8% of patients, and EVT to 84.1%, achieving successful recanalization (TICI ≥2b) in 72.6%. Despite treatment, disability remained substantial, with a median 3-month mRS of 3 (IQR, 1–6) and 71 deaths (35.3%).

**Table 1:** Comparison of baseline characteristics between patients with good and poor Outcomes.

|  | **Overall**<br>**(n=201)** | **Good outcome**<br>**(mRS=0-3; n=104)** | **Poor outcome**<br>**(mRS=4-6; n=97)** | **P-values** |
| --- | --- | --- | --- | --- |
| **BASELINE CHARACTERISTICS** |  |  |  |  |
| Age, y, median (IQR) | 70 (58-77) | 68 (57-77) | 72 (39-96) | 0.17 |
| Male, n (%) | 119 (59.2) | 63 (60.6) | 57 (58.8) | 0.79 |
| Baseline NIHSS, median (IQR) | 10 (6-18) | 8 (4-15) | 13 (8-22) | **<0.0001** |
| Patient with baseline NIHSS≤10, n (%) | 102 (50.7) | 65 (62.5) | 37 (38.1) | **0.0006** |
| **ETIOLOGY** |  |  |  |  |
| Cardio-embolic, n (%) | 65 (32.3) | 45 (43.3) | 20 (20.6) | **0.0006** |
| Atherosclerosis, n (%) | 50 (24.9) | 17 (16.3) | 33 (34.0) | **0.0038** |
| Dissection or undetermined, n (%) | 74 (36.8) | 37 (35.6) | 37 (38.1) | 0.71 |
| **RISK FACTORS** |  |  |  |  |
| History of previous stroke, n (%) | 31 (15.4) | 12 (11.8) | 19 (21.6) | 0.068 |
| Hypertension, n (%) | 112 (55.7) | 45 (43.7) | 67 (72.0) | **<0.0001** |
| Diabetes, n (%) | 34 (16.9) | 13 (12.6) | 21 (23.1) | 0.056 |
| Hyperlipidemia, n (%) | 61 (30.3) | 27 (26.2) | 34 (38.2) | 0.075 |
| Smoking, n (%) | 52 (25.9) | 24 (24.5) | 28 (33.7) | 0.171 |
| Coronary pathology, n (%) | 18 (8.9) | 6 (5.9) | 12 (13.5) | 0.076 |
| Known atrial fibrillation, n (%) | 27 (13.4) | 17 (18.3) | 10 (12.5) | 0.29 |
| **BASELINE IMAGING** |  |  |  |  |
| Time from onset to imaging, min, median (IQR) | 160 (116-256) | 160 (117-254) | 166 (115-256) | 0.76 |
| Stroke volume, mL, median (IQR) | 4.75 (1.53-11.37) | 2.38 (0.86-5.47) | 8.3 (3.14-24.74) | **<0.0001** |
| pc-ASPECT, median (IQR) | 6 (5-8) | 7 (6-8) | 6 (4-7) | **<0.0001** |
| Volume of disconnected fibers, mL, median (IQR) | 25.15 (12.99-39.08) | 14.43 (6.07-24.92) | 36.62 (27.69-53.73) | **<0.0001** |
| **TREATMENT** |  |  |  |  |
| Intravenous thrombolysis, n (%) | 84 (41.8) | 57 (54.8) | 27 (27.8) | **0.0001** |
| Endovascular treatment, n (%) | 169 (84.1) | 84 (85.7) | 85 (87.6) | 0.69 |
| Time from onset to recanalization, min, median (IQR) | 347 (259-474) | 330 (245-475) | 380 (266-468) | 0.29 |
| Successful recanalization (mTICI≥2b), n (%) | 146 (72.6) | 81 (95.3) | 65 (81.2) | **0.0047** |
| **OUTCOME** |  |  |  |  |
| mRS at 3 months, median (IQR) | 3 (1-6) | 1 (1-3) | 6 (5-6) | **<0.0001** |
| Mortality at 3 months, n (%) | 71 (35.3) | 0 (0) | 71 (73.2) | **<0.0001** |

All patients underwent acute MRI before therapy, at a median of 160 minutes from symptom onset (IQR, 116-256). Median infarct volume on DWI was small (4.75 mL; IQR, 1.53-11.37), as expected for posterior fossa strokes. However, the lesion frequency map revealed involvement across the full posterior circulation territory, indicating an exhaustive representation of BAO-related infarcts (**Figure-2, top row**).

**Figure 2:**
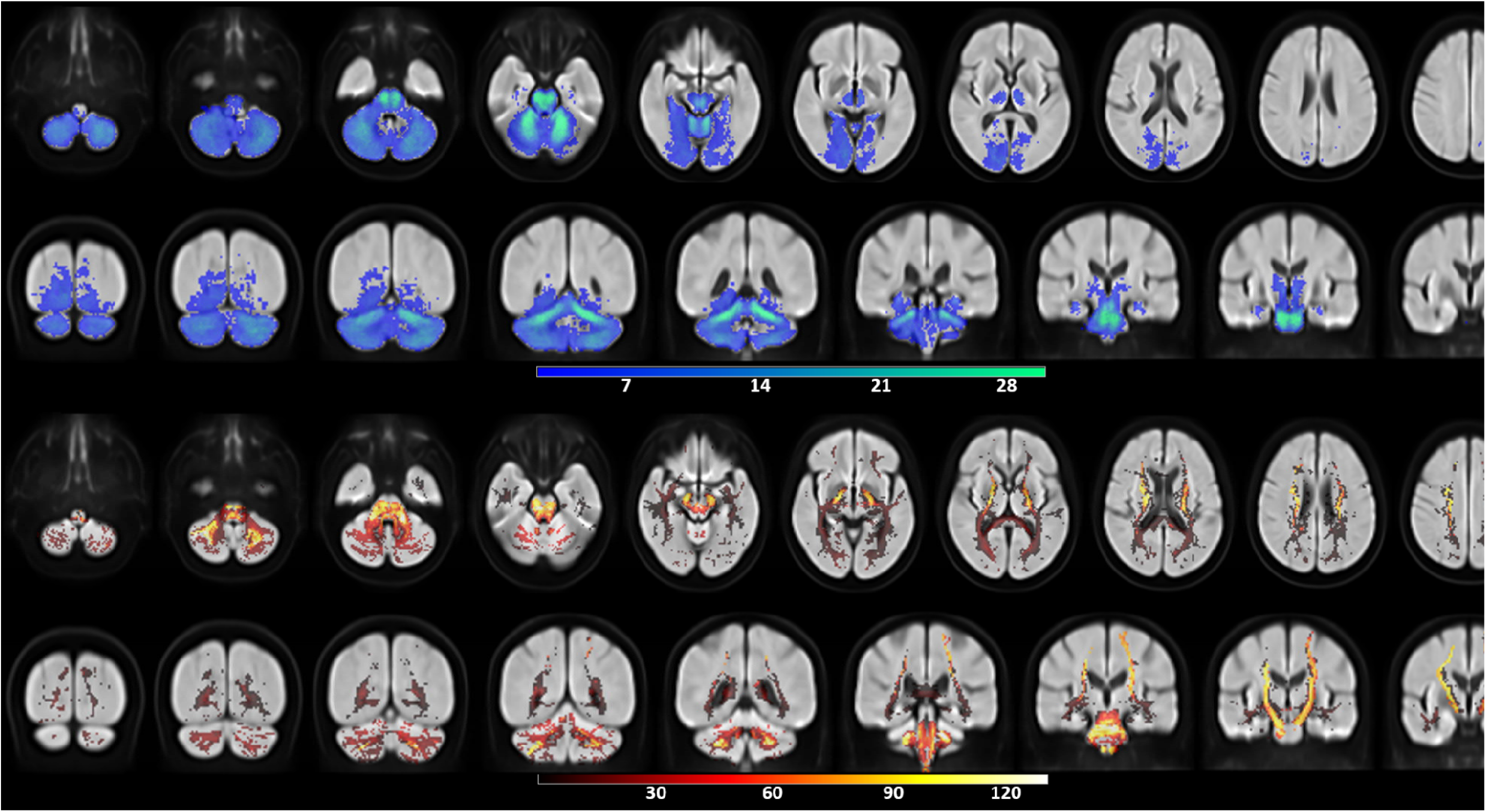
Prevalence maps of infarct lesions and associated disconnected fibers in MNI152 space. Top row: axial and coronal views showing the spatial distribution of acute infarct lesions. Bottom row: corresponding maps of disconnected white matter fibers derived from lesion projections. Color scales indicate, for each voxel, the number of patients with an infarct involving that voxel (blue scale) or with fiber disconnection passing through that voxel (red scale). The infarcts span over the entire posterior circulation territory and the disconnected fibers are more widespread.

Despite their small size, these infarcts were associated with substantially larger volumes of disconnected fibers (median, 25.15 mL; IQR, 12.99-39.08), reflecting the dense white matter architecture of the posterior fossa and its connections. Disconnections involved ascending and descending brainstem tracts, cerebellar pathways, thalamo-cortical tracts, and, depending on lesion location, the optic radiations, inferior longitudinal fasciculus, and/or forceps major of the corpus callosum. The corresponding disconnection frequency map showed a widespread distribution (**Figure-2, bottom row)**.

### Prediction of the dichotomized outcome at 3 months

At 3 months, 104 patients (51.7%) showed good outcome (mRS ≤3), while 97 (48.3%) had a poor outcome, including 71 deaths (73.2%). Compared with poor-outcome patients, those with good outcome had lower baseline NIHSS (8 [IQR, 4-15] vs. 13 [IQR, 8-22]; *p*<0.0001), more often cardioembolic stroke (43.3% vs. 20.6%; *p*=0.0006), less atherosclerosis (16.3% vs. 34.0%; *p*=0.0038), which was associated with less hypertension (43.7% vs. 72.0%; *p*<0.0001). They were more frequently treated with thrombolysis (54.8% vs. 27.8%; *p*<0.0001) and had higher rates of successful recanalization (95.3% vs. 81.2%; *p*=0.0047).

On imaging, good-outcome patients had smaller infarct volumes (*p*<0.0001), higher pc-ASPECTS (*p*<0.0001), and significantly smaller disconnected fiber volumes (*p*<0.0001; **Table 1**).

Using logistic regression, we computed and compared the predictive performances of the variables that can be available prior to EVT decision. We found that pre-treatment clinical metrics provided modest discrimination, with AUCs in the 0.6-0.7 range for baseline NIHSS (0.67 [95% CI, 0.60-0.74]) and thrombolysis eligibility (0.63 [95% CI, 0.56-0.71]; **Figure-3**). Standard imaging metrics achieved fair discrimination, with AUCs in the 0.7-0.8 range for infarct volume (0.75 [95% CI, 0.69-0.82]) and pc-ASPECTS (0.76 [95% CI, 0.69-0.82]). In contrast, disconnected fiber volume achieved a significantly higher AUC of 0.84 (95% CI, 0.78-0.89) (**Figure-3**), outperforming, baseline NIHSS (*p*<0.0001), infarct volume (*p*=0.00059) and pc-ASPECTS (*p*=0.0127).

**Figure 3:**
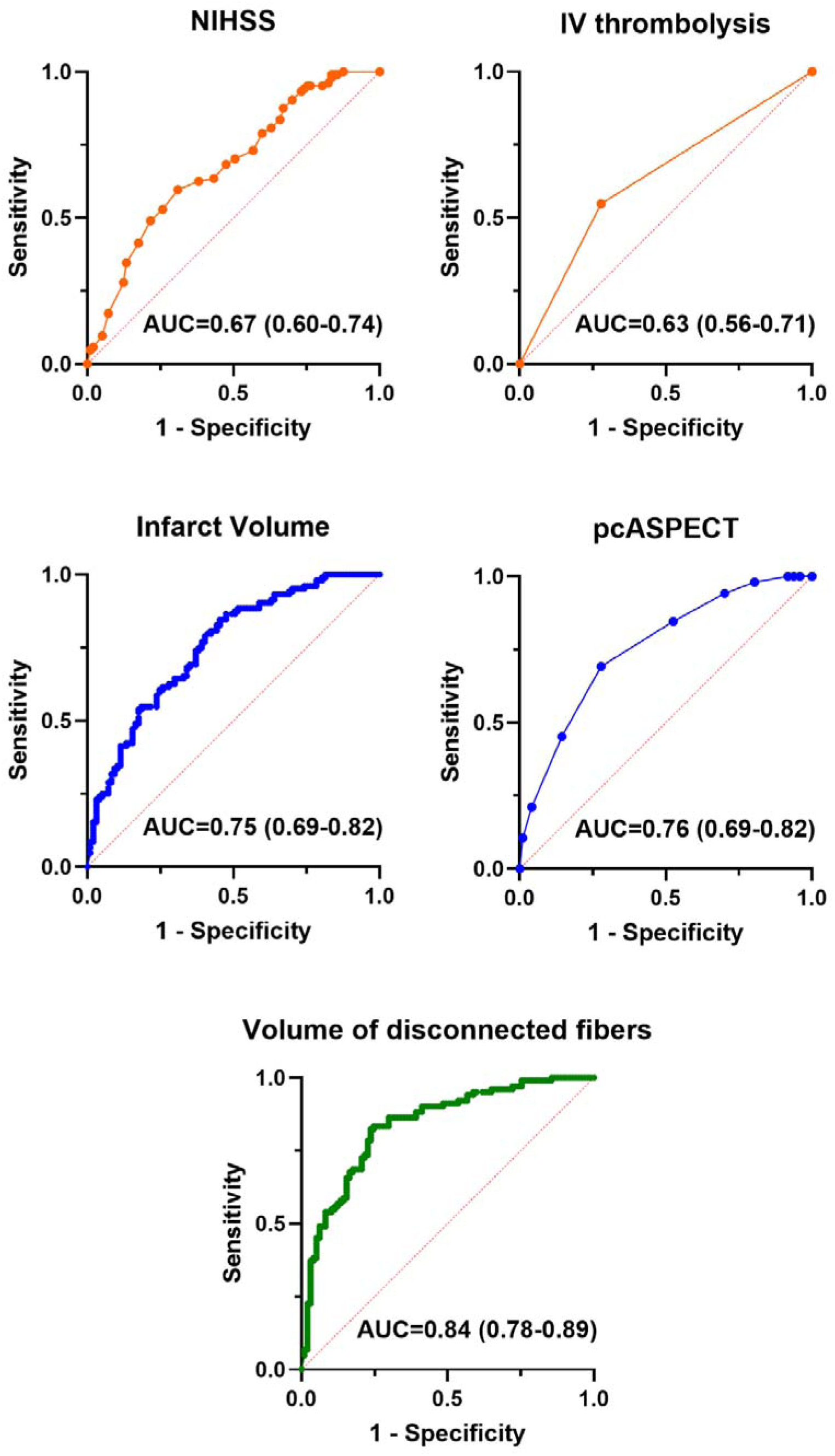
Receiver operating characteristic (ROC) curves for logistic regression models predicting dichotomized 3-month modified Rankin Scale (mRS) outcome. Curves show the area under the ROC curve (AUC) with 95% confidence intervals for the clinical predictors in orange (baseline NIHSS and eligibility for intravenous thrombolysis), the standard imaging predictors in blue (infarct volume and pc-ASPECTS), and the network-based predictor in green (volume of disconnected fibers). Prediction increased progressively from clinical predictors to standard imaging predictors up to the best accuracy for the volume of disconnected fibers.

Classification tables (probability threshold 0.5) showed correct outcome prediction in 68.6% of patients using infarct volume, 70.6% using pc-ASPECTS, and 79.4% using disconnected fiber volume. We also used a high-certainty approach defining that only patients with a predicted probability >0.7 or <0.3 could be considered at high risk or low risk to evolve toward a good outcome or poor outcome, while patients with a predicted probability of 0.3-0.7 would have an uncertain outcome. Then, replacing infarct volume with disconnected fiber volume reclassified 47.7% (96/201) of previously uncertain patients, of whom 83.3% (80/96) were correctly classified. Replacing pc-ASPECTS with disconnected fiber volume reclassified 28.8% (58/201) of uncertain patients, of whom 81.0% (47/58) were correctly classifications.

Infarct volume correlated with disconnected fiber volume, but with substantial variability for small infarcts. In patients with infarct volume <20 mL (83% of the cohort), volume explained only 33% of the variance in disconnected fibers (R²=0.33). Similarly, in those with pc-ASPECTS >3 (89% of the cohort), pc-ASPECTS explained only 24% of the variance in disconnected fibers (R²=0.24) (**Figure-4**).

**Figure 4:**
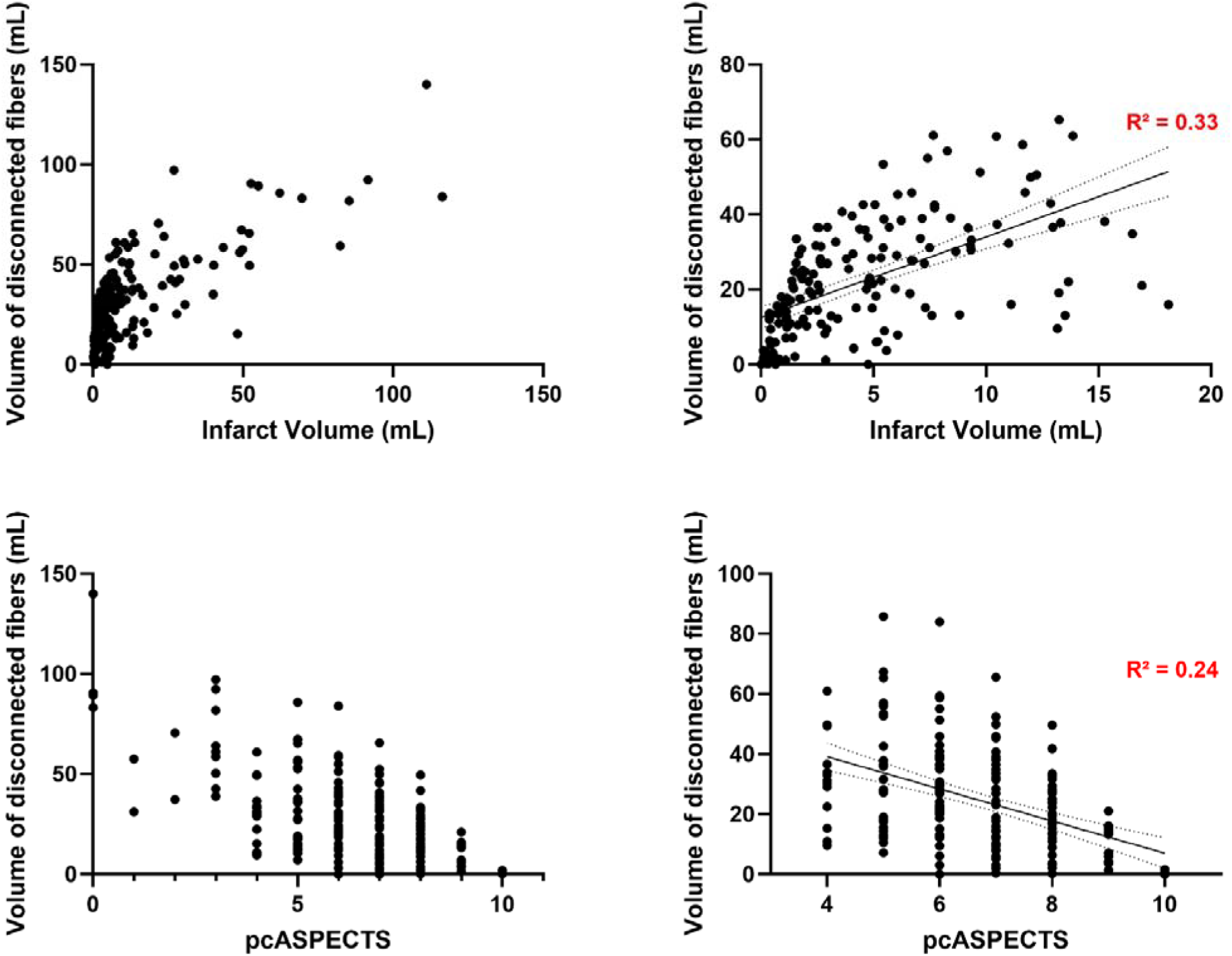
Associations between disconnected fiber volume and conventional imaging metrics. Top row: relationship between disconnected fiber volume and infarct volume. Bottom row: relationship between disconnected fiber volume and pc-ASPECTS. Plots are shown for the entire cohort and for the subgroup with smaller infarcts (volume <20 mL and pc-ASPECTS >3). In the latter subgroup, linear regression lines with 95% confidence intervals are displayed. Although infarct volume and pc-ASPECTS correlate with disconnected fiber volume, substantial variability exists, particularly in small strokes, resulting in a limited proportion of variance explained.

Collectively, these data indicate that the volume of disconnected fibers captures a new information that provides significantly higher predictive performances compared to other predictors as shown in illustrative examples in **Figure 5**.

**Figure 5:**
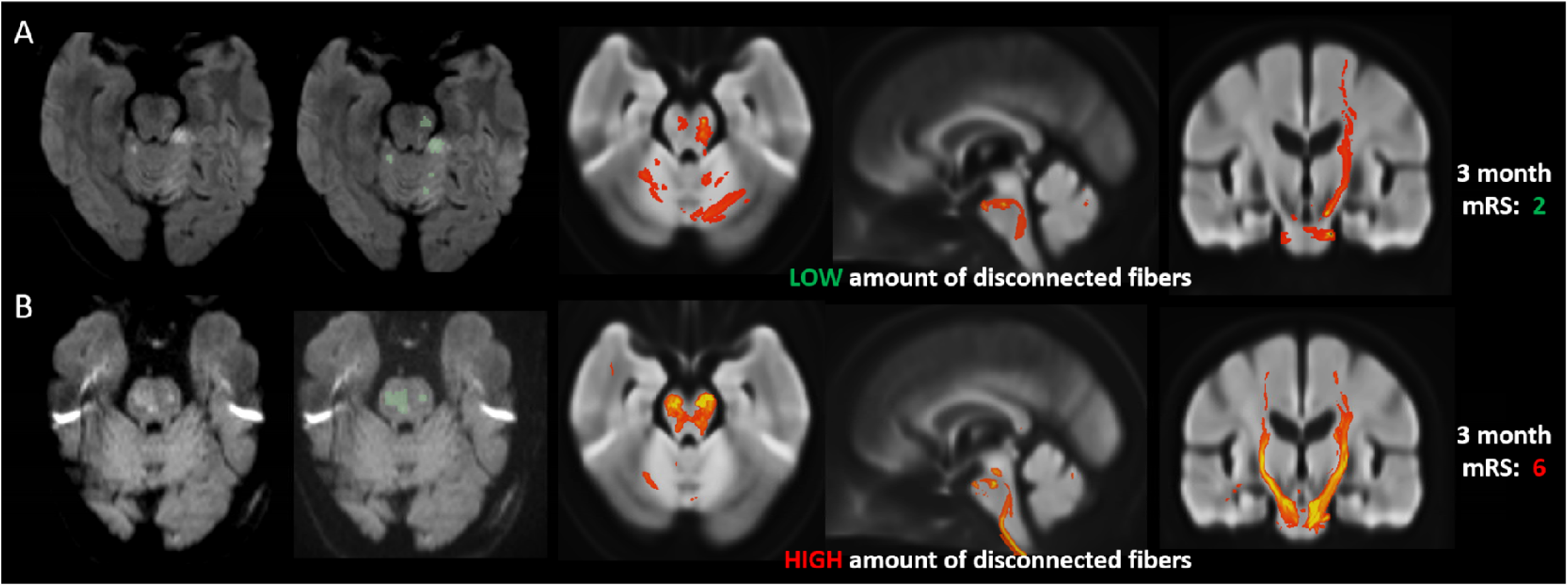
Illustrative cases of acute infarcts and corresponding disconnected fiber maps. Diffusion MRI is displayed in native space without and with manual infarct delineation (green masks). Corresponding disconnected fibers are shown in orthogonal views in MNI152 space (red). Top row is a woman in her late sixth decade with baseline NIHSS quoted at 3, a low infarct volume (3.4 mL) throughout different locations (pcASPECTS at 6) and a low volume of disconnected fibers (12.2 mL). The outcome was good with a 3-month mRS at 2. Bottom row is a woman in her seventh decade with baseline NIHSS quoted at 9, a low infarct volume as well (5.1 mL) throughout different locations (pcASPECTS at 5) but, this tiime, responsible for a high volume of disconnected fibers (42.6 mL). The outcome was poor as the patient died before 3 month.

### Prediction of Rankin shift at 3 months and interaction with recanalization status

Ordinal regression across the full mRS range demonstrated a significant shift toward better outcomes in patients with low disconnected fiber volumes at baseline (global OR=0.12 [95% CI, 0.065-0.204]; *p*<0.001) (**Figure-6**) indicating a robust predictive role of disconnected fibers whatever the mRS category.

**Figure 6:**
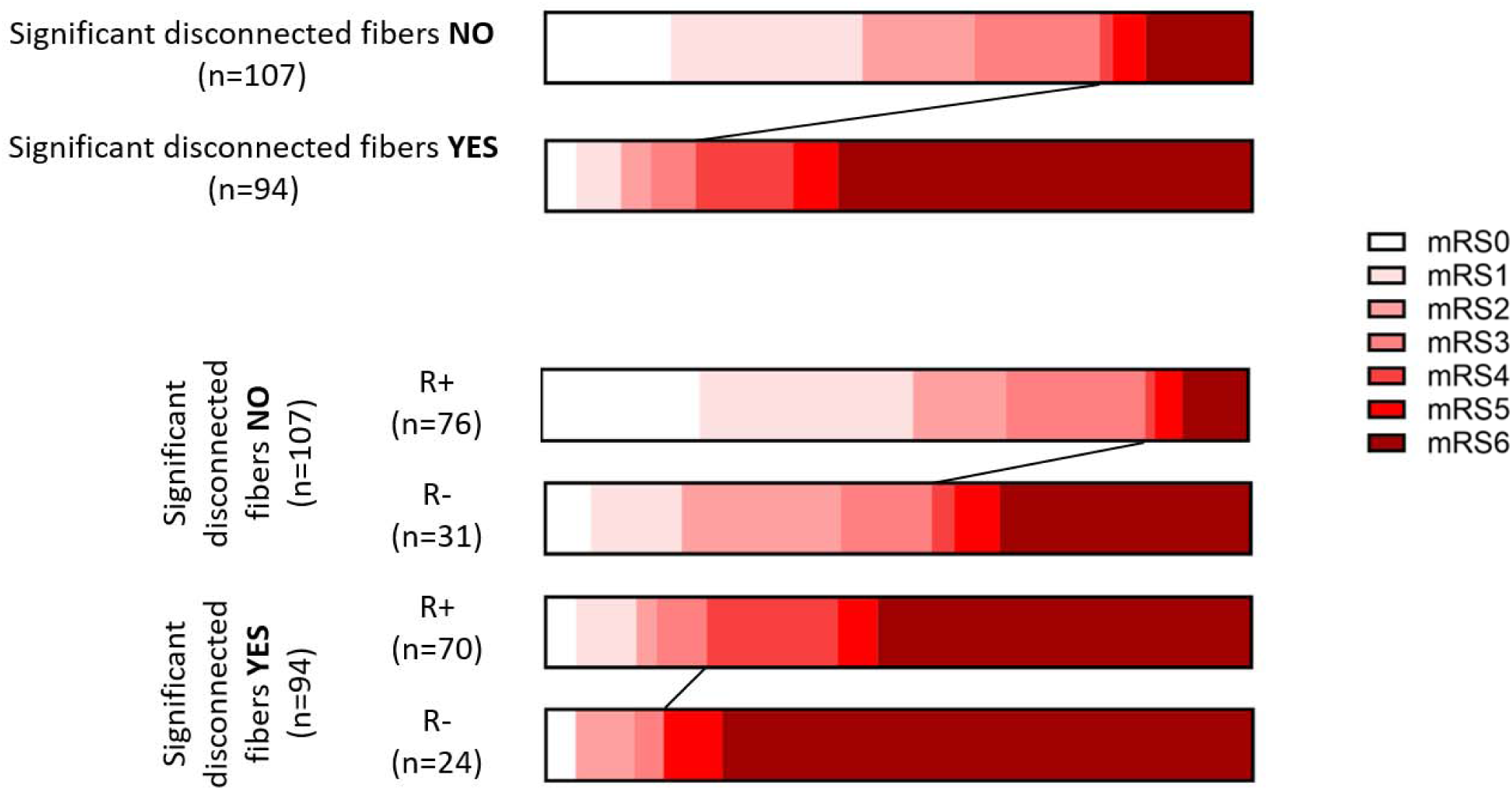
Distribution of 3 month modified Rankin Scale (mRS) scores stratified by disconnected fiber volume and recanalization status. Top row: mRS distribution according to disconnected fiber volume (low vs. high). Bottom row: mRS distribution according to disconnected fiber volume and recanalization status. Successful recanalization (**R+**) was defined as Thrombolysis in Cerebral Infarction (TICI) ≥2b; unsuccessful recanalization (**R−**) as TICI 0–2a. Oblique lines highlight the shift in outcome for mRS ≤3. Baseline disconnected fiber volume was associated with the distribution of mRS scores at 3 months and appeared to influence the benefit from recanalization.

In the low-disconnection group, successful recanalization was associated with a significant and substantial shift toward better 3-month mRS (OR=0.33 [95% CI, 0.15-0.70]). In the high-disconnection group, recanalization showed a trend toward benefit, but the effect was smaller and not statistically significant (OR=0.39 [95% CI, 0.14-1.09]) (**Figure-6**).

### Secondary analyses in patients with mild baseline deficit

Given the uncertainty of reperfusion benefit in BAO patients with mild initial deficits ^4, 5^, we repeated the analyses in the subgroup with baseline NIHSS ≤10 (n=102; 50.7%) to understand whether the volume of disconnected fibers from acute MRI could provide some guidance for this category of patients. In this subgroup, disconnected fiber volume remained the strongest predictor of dichotomized outcome (AUC=0.83 [95% CI, 0.74-0.92]) (**Supplementary-Figure-2**).

Ordinal regression again showed a marked shift toward better outcomes in the low-disconnection group (global OR=0.089 [95% CI, 0.036-0.224]; *p*<0.00001) (**Supplementary-Figure-3**). The sample size was insufficient to stratify these results by recanalization status.

## Discussion

In this multicenter study, we showed that the volume of disconnected white matter fibers, estimated automatically from acute DWI lesions without requiring patient-specific diffusion tractography, is a robust prognostic marker of 90-day functional outcome in acute BAO, outperforming conventional predictors. This metric also identified subgroups more likely to benefit from EVT, even in patients with moderate baseline severity, highlighting its potential for imaging-guided therapeutic decision-making and for selecting targeted populations in future neuroprotective trials.

BAO remains a devastating condition ^1^. In our cohort, approximately half of the patients were non-ambulatory at 3 months and more than one third died, despite a high rate of EVT, findings consistent with previous reports ^2, 3, 5, 23^. The decision to pursue EVT can be particularly challenging in clinical practice, for example, when long transfers to comprehensive stroke centers are required, in frail or elderly patients, or in those with mild initial deficits as there is no clear evidence of EVT benefit when NIHSS <10 ^4, 5^. Prognostic tools could therefore be invaluable, not only to guide treatment selection, but also to inform families and plan home adaptations rapidly ^34^. Such tools could also identify more homogeneous patient populations to tailor new neuroprotective strategies to specific patients in future trials, thereby improving statistical power ^35, 36^. Our findings show that accurate prognostication is achievable using acute imaging acquired before any treatment.

Whereas outcome prediction in anterior circulation stroke is strongly influenced by age and baseline NIHSS ^34, 37^, these metrics are less useful in BAO, partly because the NIHSS is weighted toward anterior circulation deficits ^8^. In our study, stroke volume and pc-ASPECTS improved predictive accuracy compared to the clinical metrics, but both fell short of the critical 0.8 AUC threshold. Several alternative imaging-based posterior circulation scoring systems have been proposed, including the Brainstem Score (BSS) ^38^, Pons-Midbrain-Thalamus (PMT) score ^11^, and Critical Area Perfusion Score (CAPS) ^39^ among others. But these share similar limitations with pc-ASPECTS: they rely on semi-quantitative location-based assessment, assume segregated rather than network-based brain function, and are non-continuous measures. Moreover, most have been validated in relatively small cohorts.

The volume of disconnected fibers addresses these limitations by providing a continuous, network-based metric that captures the disruption of structural brain interactions. Our findings extend prior work in anterior circulation stroke, where disconnectome approaches have shown potential in predicting complex outcomes such as post-stroke cognition ^14, 17, 18^. To our knowledge, this is the first application of this method to posterior circulation stroke, demonstrating its added value for global prognostication, likely reflecting the critical role of brainstem and cerebellar networks in motor, sensory, and arousal systems. This was possible thanks to dedicated optimization of the pipeline to address specific posterior fossa challenges. We first finely tuned the co-registration, mitigating substantial EPI-related geometric distortions caused by adjacent air-filled structures, as even millimeter-scale misalignment can significantly alter disconnection estimates in the ultra-dense brainstem network. We also improved tractography quality, acquiring ultra–high-resolution diffusion MRI (10-hour acquisitions) to reliably resolve complex posterior fossa fiber bundles. These methodological advances yielded the disconnected fiber volume, a metric only partly explained by infarct volume or location, yet strongly linked to functional outcome and to the likelihood of benefit from recanalization.

Still, some limitations warrant consideration. First, external validation is needed to confirm generalizability. Nevertheless, our large multicenter dataset included lesions distributed throughout the entire posterior circulation territory already supports heterogeneity and representativeness. Second, disconnectome analysis is indirect and does not capture individual variability in structural connectivity. However, it uniquely enables network-level assessment in the acute setting, when there is no time for individualized multi-direction diffusion imaging ^40^, by leveraging prior knowledge from ultra–high-resolution datasets. Third, the current workflow is not real-time, requiring lesion delineation, co-registration, and projection steps. We have already developed a supra-tentorial disconnectivity toolkit ^16^, and a posterior fossa-specific version is feasible. We have also recently proposed a deep-learning version of the disconnectivity tool ^41^ to efficiently speed up the pipeline which we plan to develop for the posterior fossa as well with the final objective to offer real-time support for therapeutic decision-making. Finally, this approach requires acute MRI, which is not universally available. With MRI increasingly integrated into emergency departments and most implants now proven MRI-safe ^42^, more comprehensive stroke centers may consider MRI as a first-line modality ^43^. Even though, this might not be feasible to get acute MRI for any stroke, this could be indicated at least for the selected BAO patients.

Overall, incorporating structural connectivity into prognostic models moves beyond traditional topographic approaches, enabling more sensitive and individualized predictions. In BAO, where lesion size, location, and presentation vary widely, disconnectome analysis refines outcome prediction and may help identify patients most likely to benefit from EVT. Future prospective studies and the development of automated, user-friendly clinical tools will be essential to confirm these findings and to evaluate their impact on treatment decisions in routine practice.

## Abbreviations

ADC: Apparent Diffusion Coefficient
ANTs: Advanced Normalization Tools
AUC: Area Under the receiver operating characteristic Curve
BAO: Basilar Artery Occlusion
DWI: Diffusion Weighted Imaging
ETIS registry: Endovascular Treatment in Ischemic Stroke registry
EVT: Endovascular Treatment
FOD: Fiber Orientation Distribution
MNI space: Montreal Neurological Institute space
mRS: modified Rankin Scale
NIHSS: National Institutes of Health Stroke Scale
pcASPECTS: posterior circulation Alberta Stroke Program Early Computed Tomography Score
TICI: Thrombolysis in Cerebral Infarction

## Data availability

Data could be made available upon request to the principal investigators.

## Supplementary material

**Supplementary Figure 1:**
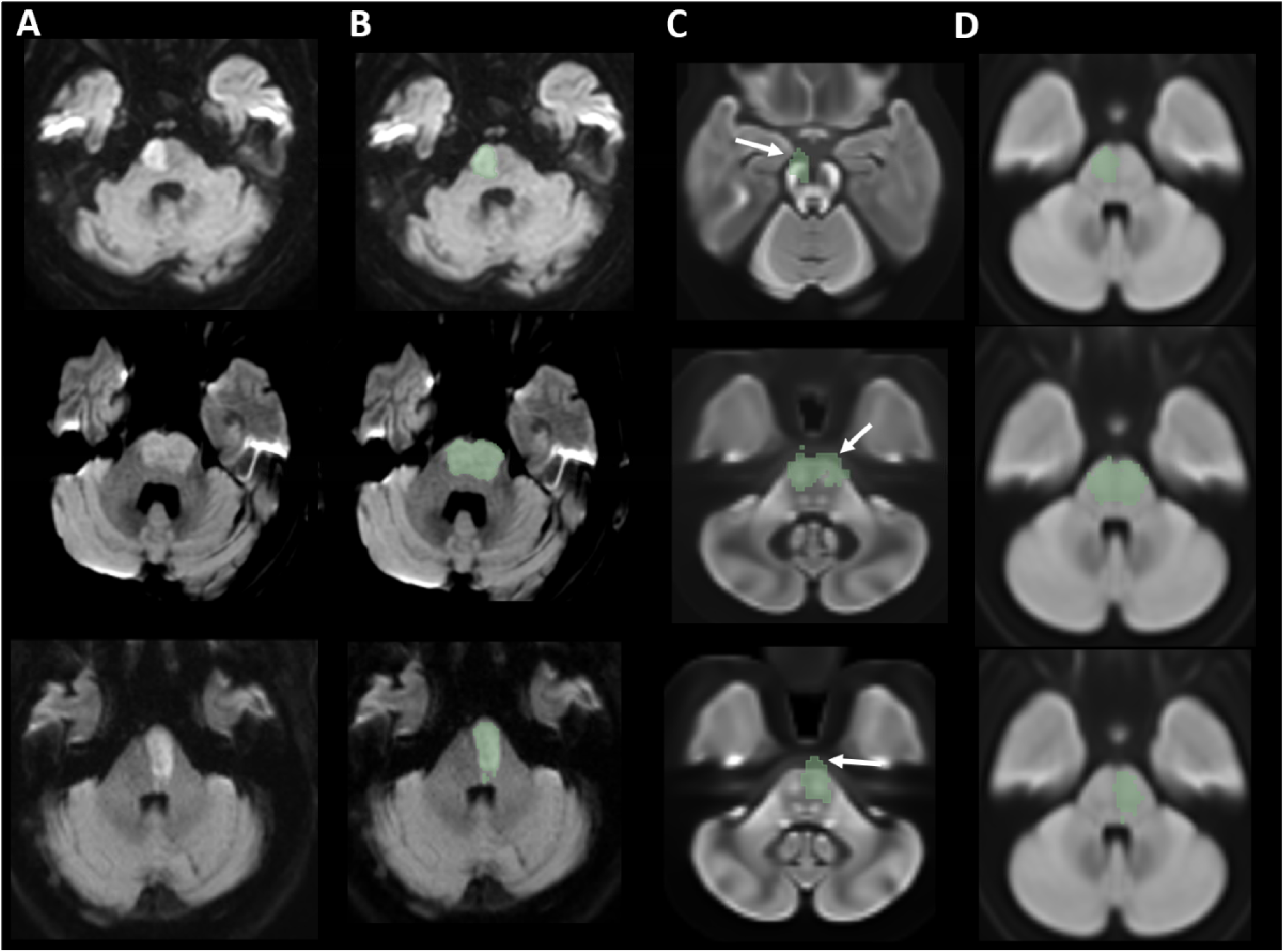
Illustration of the improved coregistration pipeline in three representative cases. (**A**) Diffusion MRI in native space. (**B**) Same images with manual infarct delineation (green masks). (**C**) Coregistration to MNI152 space using ANTs, showing misalignment (white arrows) due to imperfect correction of brainstem geometric distortions. (**D**) Coregistration accuracy after incorporating an intermediate step with a study-specific DWI template, demonstrating substantial improvement.

**Supplementary Figure 2:**
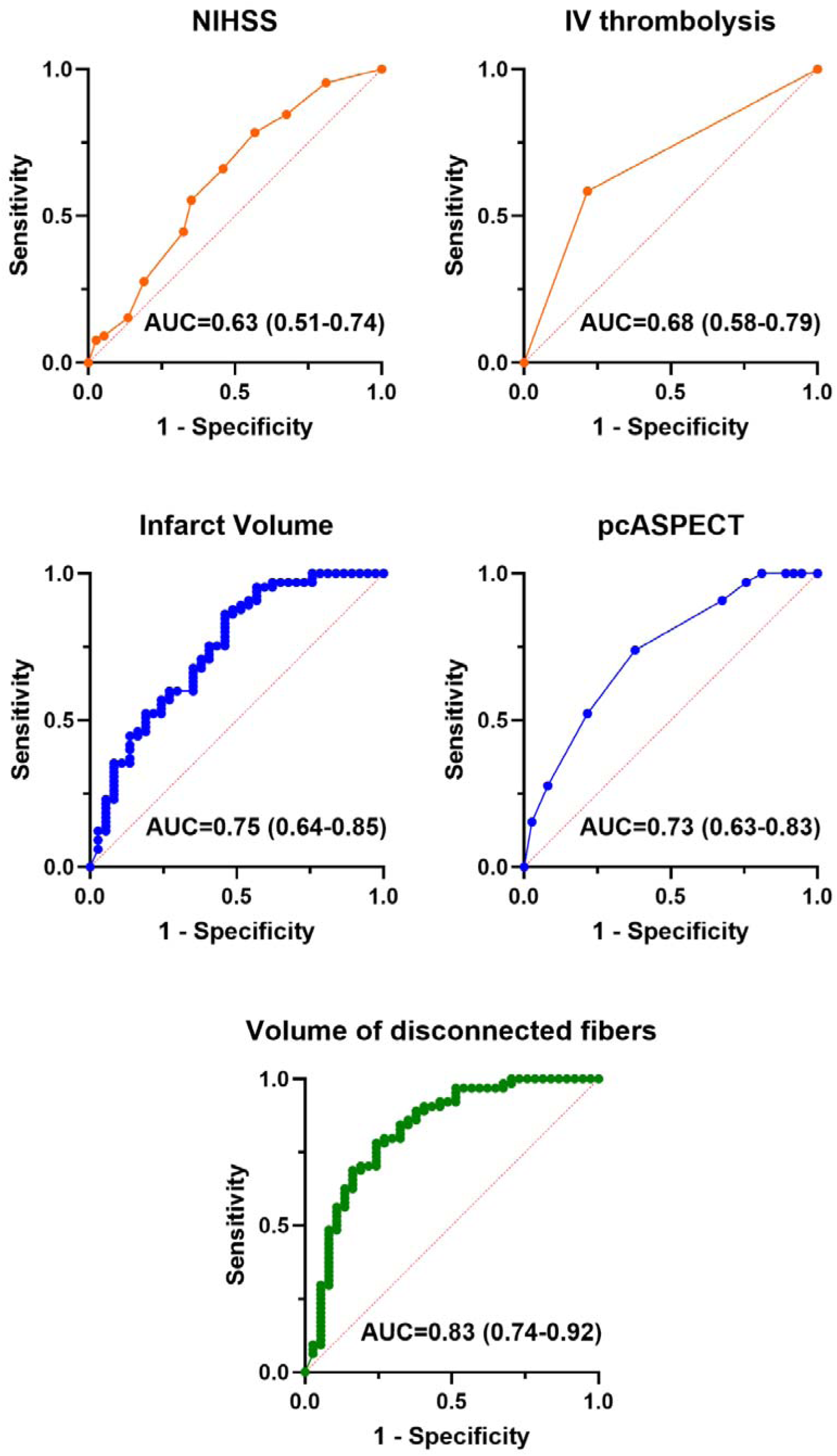
Receiver operating characteristic (ROC) curves for logistic regression models predicting dichotomized 3-month modified Rankin Scale (mRS) outcome for the subgroup of patients with mild initial deficit defined as baseline NIHSS ≤10. The data are represented for the 102 patients with baseline NIHSS≤10. Curves show the area under the ROC curve (AUC) with 95% confidence intervals for the clinical predictors in orange (baseline NIHSS and eligibility for intravenous thrombolysis), the standard imaging predictors in blue (infarct volume and pc-ASPECTS), and the network-based predictor in green (volume of disconnected fibers). Prediction increased progressively from clinical predictors to standard imaging predictors up to the best accuracy for the volume of disconnected fibers.

**Supplementary Figure 3:**
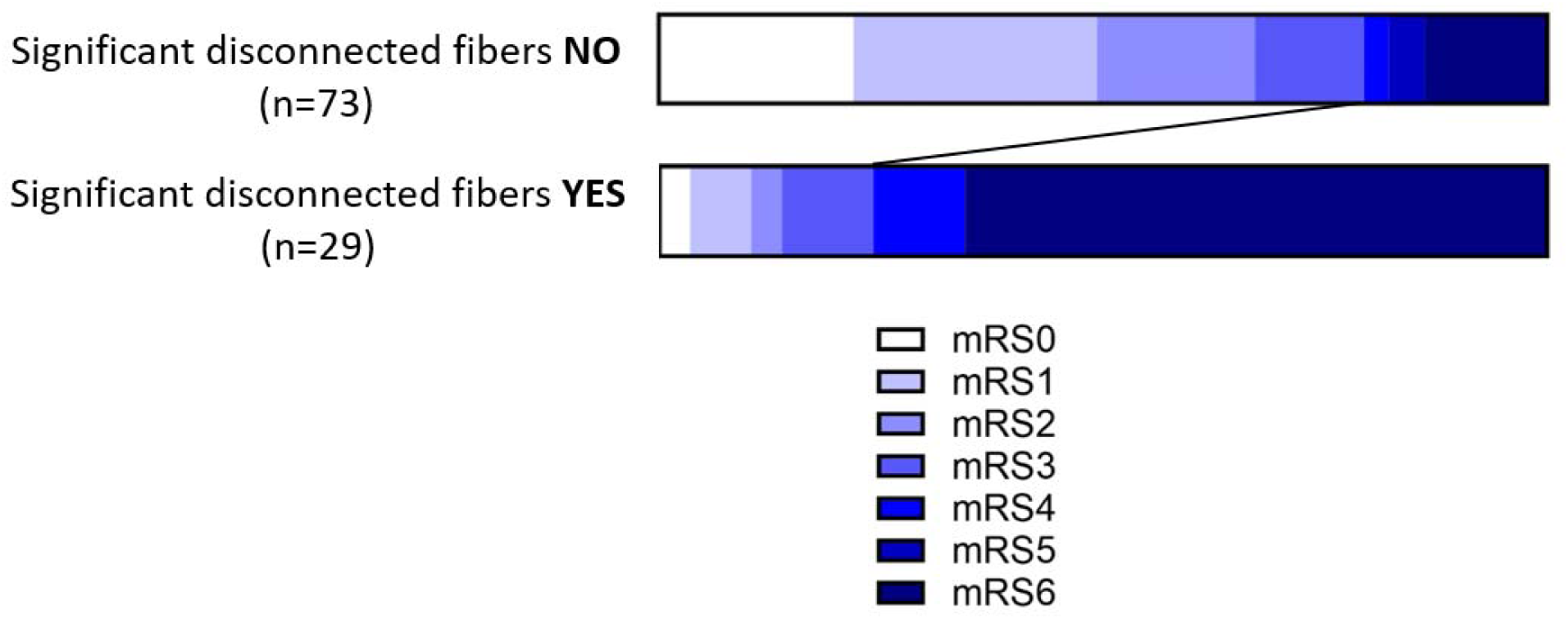
Distribution of 3 month modified Rankin Scale (mRS) scores stratified by disconnected fiber volume for the subgroup of patients with mild initial deficit defined as baseline NIHSS ≤10 (n=102). Oblique line highlights the shift in outcome for mRS ≤3.

